# *In vivo* NMDA receptor function in people with NMDA receptor antibody encephalitis

**DOI:** 10.1101/2021.12.04.21267226

**Authors:** Marian Galovic, Adam Al-Diwani, Umesh Vivekananda, Francisco Torrealdea, Kjell Erlandsson, Tim D Fryer, Young T Hong, Benjamin A Thomas, Colm J McGinnity, Evan Edmond, Kerstin Sander, Erik Årstad, Ilijas Jelcic, Franklin I Aigbirhio, Ashley M Groves, Kris Thielemans, Brian Hutton, Alexander Hammers, John S Duncan, Jonathan P Coles, Anna Barnes, Charlotte J Stagg, Matthew C Walker, Sarosh R Irani, Matthias J Koepp, for the NEST investigators

**Affiliations:** Department of Neurology, Clinical Neuroscience Center, University Hospital Zurich, Zurich, Switzerland; Department of Clinical and Experimental Epilepsy, UCL Queen Square Institute of Neurology, University College London, London, UK; MRI Unit, Chalfont Centre for Epilepsy, UK; Oxford Autoimmune Neurology Group, Nuffield Department of Clinical Neurosciences, University of Oxford, Oxford, United Kingdom; Department of Psychiatry, University of Oxford, Oxford, United Kingdom; Department of Medical Physics and Biomedical Engineering, University College London Hospitals, London, UK; Institute of Nuclear Medicine, University College London Hospitals, London, UK; Wolfson Brain Imaging Centre, Department of Clinical Neurosciences, University of Cambridge, Cambridge, UK; King’s College London & Guy’s and St Thomas’ PET Centre, School of Biomedical Engineering and Imaging Sciences, King’s College London, London, UK; Wellcome Centre for Integrative Neuroimaging & Medical Research Council Brain Network Dynamics Unit, Nuffield Department of Clinical Neurosciences, University of Oxford, Oxford, United Kingdom; Centre for Radiopharmaceutical Chemistry, University College London, London, UK; Division of Anaesthesia, Department of Medicine, University of Cambridge, Cambridge, UK; Department of Neurology, John Radcliffe Hospital, Oxford University Hospitals NHS Foundation Trust, Oxford, UK

**Author notes:** Corresponding author: Prof. Matthias Koepp, UCL Institute of Neurology, Queen Square, London WC1N 3BG. Full list of NEST Investigators is given in the online supplement.

## Abstract

In *N*-methyl-D-aspartate receptor (NMDAR) antibody encephalitis, NMDAR-autoantibodies are hypothesised to cause prominent neuropsychiatric symptoms by internalizing NMDARs. However, supporting evidence comes chiefly from *in vitro* and rodent data with scant direct evidence from affected humans. Here, we used *in vivo* positron emission tomography (PET) with [^18^F]GE-179 to show a mean 30% reduction of the density of open, activated NMDARs in grey matter of persistently NMDAR-autoantibody seropositive patients following NMDAR-antibody encephalitis compared to healthy controls. The reduction was most prominent in the anterior temporal and superior parietal cortices. These patients had normal structural MRIs and mild residual symptoms. In contrast, one symptom-free patient who recovered from NMDAR-antibody encephalitis and was not NMDAR-autoantibody seropositive had normal density of active NMDARs. These findings reveal a functional deficit of open, activated NMDARs in humans with NMDAR-autoantibodies. Moreover, we observed a functional NMDAR deficit for up to 8 months following the disease peak, despite only mild residual symptoms, highlighting the considerable compensatory capacity of the human brain.

**One Sentence Summary:** Reductions of activated NMDA receptors detected *in vivo* in female patients following NMDA-receptor-antibody encephalitis.

## INTRODUCTION

*N*-methyl-D-aspartate receptor (NMDAR)-autoantibodies directed against the GluN1 subunit of the NMDAR mediate the commonest established form of autoimmune encephalitis. It presents with neuropsychiatric features alongside seizures, speech abnormalities, movement disorders, and a decreased level of consciousness *(1, 2)*. Around 80% of cases have a favourable outcome following immunotherapy. However, relapses may occur in up to ∼25% *(3, 4)* and the majority of recovered cases continue to report persistent cognitive deficits, particularly affecting executive functions and memory *(5)*. These persistent symptoms are refractory to immunotherapies, and the underlying mechanisms remain unclear.

*In vitro* studies indicate that NMDAR-autoantibodies crosslink and internalise NMDARs, possibly through disruption of the NMDAR interaction with Ephrin-B2 receptors *(6, 7)*. A selective loss of NMDARs, without disruption of other synaptic components, is also observed after passive transfer of NMDAR Immunoglobulin G (IgG) to experimental animals *(8, 9)*.

Together, these observations have led to the hypothesis that internalisation of NMDAR is a major pathogenic mechanism underlying NMDAR-antibody encephalitis. However, direct support for this in humans is limited to a report showing reduction of NMDAR staining in the *post mortem* hippocampi of two autopsied patients *(9). In vivo* studies of humans with NMDAR-antibody encephalitis have examined patients using magnetic resonance imaging (MRI) *(10, 11)* and [^18^F]fluorodeoxyglucose (FDG) positron emission tomography (PET) *(12)* but none have directly measured NMDAR activity.

We performed a cross-sectional functional imaging study of NMDARs *in vivo* in five patients during recovery from definite NMDAR-antibody encephalitis *(13)*, using PET with the radioligand [^18^F]GE-179. This compound binds within the ion channel of the open, i.e. activated, NMDAR. Recent *in vivo* blocking experiments performed following electrical stimulation designed to activate NMDARs showed the specificity of [^18^F]GE-179 for the phencyclidine site as a use-dependent marker of NMDAR activation *(14)*. In contrast, blocking experiments in anaesthetised animals failed *(15)*, potentially due to a reduced baseline opening probability of NMDARs following anaesthesia *(16)*. This provides evidence that the radioligand is sensitive to manipulations that activate NMDAR. We used [^18^F]GE-179 PET to determine whether disturbances of NMDAR function improve after recovery from acute NMDAR-antibody encephalitis and whether they correlate with NMDAR-autoantibody titres, with [^18^F]GE-179 total volume of distribution (V_T_) used to quantify open, activated NMDAR density.

## RESULTS

### [18F]GE-179 PET binding is reduced in seropositive females following NMDAR-antibody encephalitis

Five patients (mean age 28 ± 5 years, all female) with definite NMDAR-antibody encephalitis *(13)* were imaged at University College London 2 to 16 months from hospital discharge. Four patients (cases #1-4) who had persistent GluN1-autoantibodies in serum (titre range 1:160 to 1:320) and were scanned two to eight months after hospital discharge were classified as persistently “seropositive”. At the time of PET imaging, these four patients had mild symptoms. One completely symptom-free patient (case #5) who no longer had serum GluN1-antibodies was scanned 14 to 18 months after discharge from hospital, and was classified as “seroreverted”. The clinical MRI was normal in all patients. Patient characteristics are displayed in **Table 1**. For comparison, we included 29 healthy volunteers (mean age 41 ± 13 years, 8 [28%] female; 10 scanned at UCL, 19 at two additional sites).

**Table 1:** Characteristics of patients with NMDAR-antibody encephalitis

| Variable | Case #1 | Case #2 | Case #3 | Case #4 | Case #5 |
| --- | --- | --- | --- | --- | --- |
| **Age-range, Sex** | 21-25, F | 36-40, F | 21-25, F | 26-30, F | 26-30, F |
| **Data at scan** |  |  |  |  |  |
| GE-179 V<sub>T</sub> in GM | 7.3 | 6.6 | 6.0 | 4.6 | 9.7 |
| Serum GluN1-IgG | 1:320 | 1:320 | 1:160 | 1:320 | Not detectable |
| Current episode | Relapse | Relapse | First | First | First |
| Overall number of episodes | 3 | 2 | 1 | 1 | 1 |
| ACE-III | 96 | 95 | 86 | 96 | 96 |
| BDI | 16 | 3 | 8 | 6 | 5 |
| **Timing** |  |  |  |  |  |
| Time from discharge to scan (months) | 2 | 8 | 6 | 8 | 16 |
| Duration of hospitalization (months) | 1.5 | 1.3 | 10 | 2.5 | 2 |
| Time from episode onset to admission (days) | 14 | 21 | 64 | 12 | 25 |
| Time from episode onset to scan (months) | 4 | 12 | 18 | 11 | 19 |
| **Treatment** |  |  |  |  |  |
| Medication at time of PET scan | CP, Prednisone, Lorazepam, TMP/SMX, Omeprazole | Levetiracetam, Clonazepam, Ferrous fumarate | Prednisone, Lansoprazole | Levetiracetam | Oral contraceptive |
| Immunotherapy during hospitalisation | Steroids, PLEX, CP | Steroids, PLEX, CP | Steroids, PLEX, IVIG | Steroids, PLEX | Steroids, PLEX, IVIG, CP |
| Other treatments during hospitalisation | Antipsychotic, Benzodiazepine | Antipsychotic, Antiseizure, Benzodiazepine | Antipsychotic, Antiseizure, Benzodiazepine | Antiseizure | Antipsychotic, Antiseizure, Benzodiazepine |
| **Clinical features during hospitalisation** |  |  |  |  |  |
| Hospitalised in | Oxford | Oxford | London | London | Oxford |
| CASE score at discharge | 5 | 4 | 5 | 5 | 8 |
| Prodrome | Y | Y | Y | Y | Y |
| Psychiatric | Y | Y | Y | Y | Y |
| Cognitive | Y | Y | Y | Y | Y |
| Speech dysfunction | Y | Y | N | Y | Y |
| Seizures | N | N | Y | Y | Y |
| Movement disorder | Y | Y | Y | Y | Y |
| Reduced consciousness | N | N | Y | Y | N |
| Autonomic | Y | N | N | N | Y |
| Central hypoventilation | N | N | N | N | N |
| Ovarian teratoma | N | N | Y | Y | N |
| Post-HSV encephalitis | N | N | N | N | N |
| **Para-clinical features during hospitalisation** |  |  |  |  |  |
| MRI abnormal | N | N | N | N | N |
| EEG abnormal | Y | Y | Y | Y | ? |
| CSF pleocytosis | Y | Y | N | N | Y |
| CSF lymphocytosis | Y | Y | Y | Y | Y |
| CSF elevated protein | Y | Y | N | N | Y |

Age ranges are presented due to confidentiality issues. Y, Yes/Present; N, No/Absent; ?, unknown; F, female; CM, Grey matter; ACE-III, Addenbrooke's Cognitive Examination III; BDI, Beck's Depression Inventory; CP, Cyclophosphamide, TMP/SMX, Trimethoprim/sulfamethoxazole; PLEX, Plasma exchange; IVIG, Intravenous immunoglobulins; HSV, Herpes simplex virus; MRI, Magnetic resonance imaging; EEG, Electroencephalography; CSF, Cerebrospinal fluid examination.

Persistently seropositive cases #1-4 had lower [^18^F]GE-179 V_T_ in comparison to the seroreverted case #5 and to healthy volunteers (**Fig. 1**). Seropositive cases had lower [^18^F]GE-179 V_T_ in grey matter (estimated marginal mean corrected for age, sex, and site: 6.2, 95% confidence interval [CI] 4.4 – 8.0) compared to healthy volunteers (8.8, 95% CI 8.1 – 9.4, F=6.5, p=0.02). We performed a voxel-based analysis to address the spatial distribution of the findings (**Fig. 2B**), which showed reduced [^18^F]GE-179 V_T_ in persistently seropositive cases within bilateral anterior temporal lobes (left, T = 4.5, 6537 mm^3^, p_FWE_ = 0.02; right, T = 4.9, 4779 mm^3^, p_FWE_=0.05) and a large cluster involving bilateral superior parietal lobes, paracentral lobules, left posterior cingulate gyrus, and left precuneus (T = 5.8, 22069 mm^3^, p_FWE_ < 0.001). *A priori* volume of interest analyses of radioligand V_T_ in each cerebral lobe corroborated the voxel-based analysis, showing approximately 30% reductions in the regional V_T_ estimates in the temporal (34%; cases 5.7 [95% CI 3.9 – 7.6] vs. controls 8.6 [95% CI 8.0 – 9.2], F=8.3, p=0.008) and parietal (31%; cases 6.2 [95% CI 4.3 – 8.2] vs. controls 9.1 [95% CI 8.4 – 9.8], F=7.3, p=0.01) lobes (**Fig. 2A**). V_T_ in white matter was comparable between seropositive cases (4.3 [95% CI 2.6 – 6.0]) and healthy volunteers (5.6 [95% CI 5.0 – 6.2], F=2.0, p=0.17 **Fig. 2A)**. Seroreverted case #5 had V_T_ estimates similar to healthy volunteers in grey (9.7) and white (5.8) matter (**Fig. 1**).

**Figure 1:**
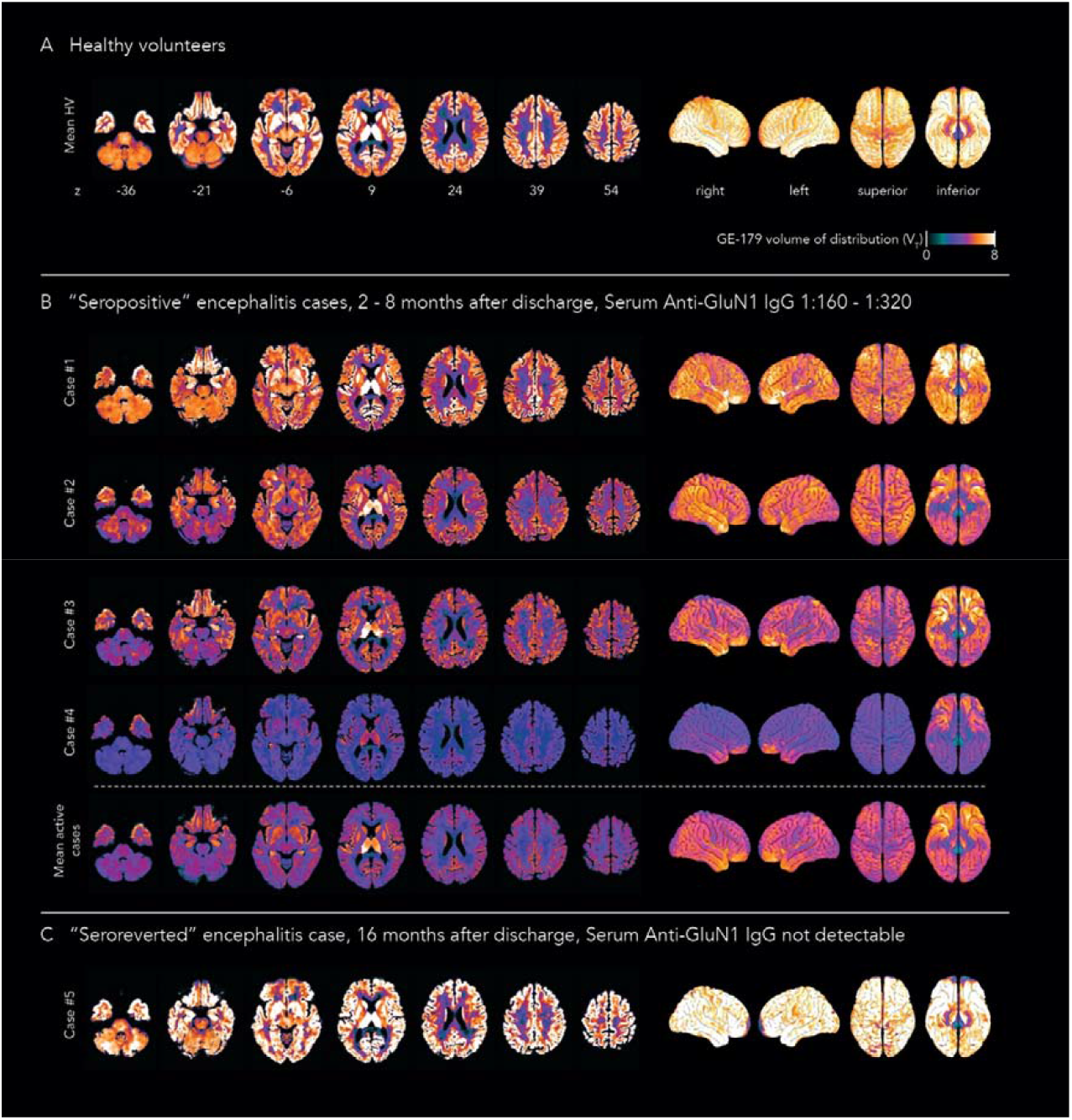
[^18^F]GE-179 uptake in autoantibody “seropositive” or “seroreverted” patients with NMDAR-antibody encephalitis and healthy volunteers. The figure shows the spatial distribution of [^18^F]GE-179 total volume of distribution (V_T_) on brain slices and surface projections. **Panel A** shows mean uptake in healthy volunteers (HV, n=29). **Panel B** displays individual V_T_ distributions in persistently autoantibody “seropositive” NMDAR-antibody encephalitis cases (n=4) that were scanned 2-8 months after discharge and had elevated serum GluN1-autoantibodies (1:160 – 1:320). The mean brain V_T_ in these cases is shown in the last row. **Panel C** displays one “seroreverted” NMDAR-antibody encephalitis case scanned 16 months after discharge with undetectable GluN1-autoantibodies on the day of scanning.

**Figure 2:**
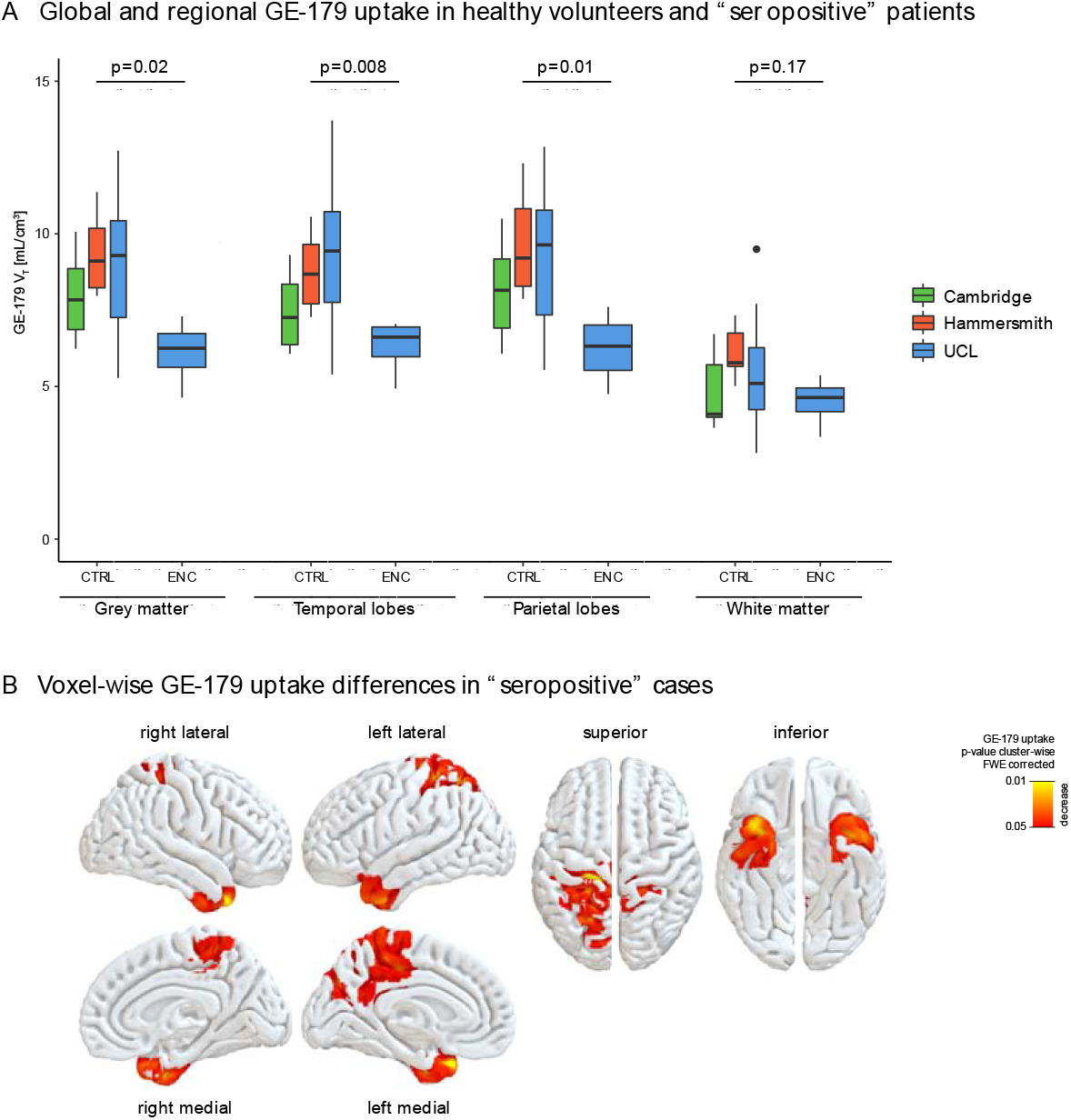
Statistical analyses of [^18^F]GE-179 uptake in NMDAR-antibody encephalitis cases and healthy volunteers. Panel **A** shows boxplots of regional [^18^F]GE-179 total volume of distribution (V_T_) in healthy volunteers (CTRL, n=29, split by cohort) and persistently autoantibody “seropositive” cases with NMDAR-antibody encephalitis (ENC, n=4). **Panel B** shows whole brain voxel-wise differences in [^18^F]GE-179 V_T_ between persistently “seropositive” encephalitis cases and healthy volunteers (p<0.05 family wise error [FWE] corrected, red colours indicate decreased uptake in encephalitis cases).

There were non-significant trends towards higher grey matter [^18^F]GE-179 V_T_ with lower serum GluN1 immunoglobulin G levels and longer time from hospital discharge and from episode onset (**Fig. 3A-C)**. Grey matter V_T_ did not correlate with cognitive testing results at the time of scanning (Addenbrooke’s Cognitive Examination III [ACE-III] questionnaire, range 86 to 96, **Fig. 3D**) nor symptom severity at discharge (CASE score at discharge, range 4 to 8, **Fig. 3E**), but there was little variability between patients in these test results.

**Figure 3:**
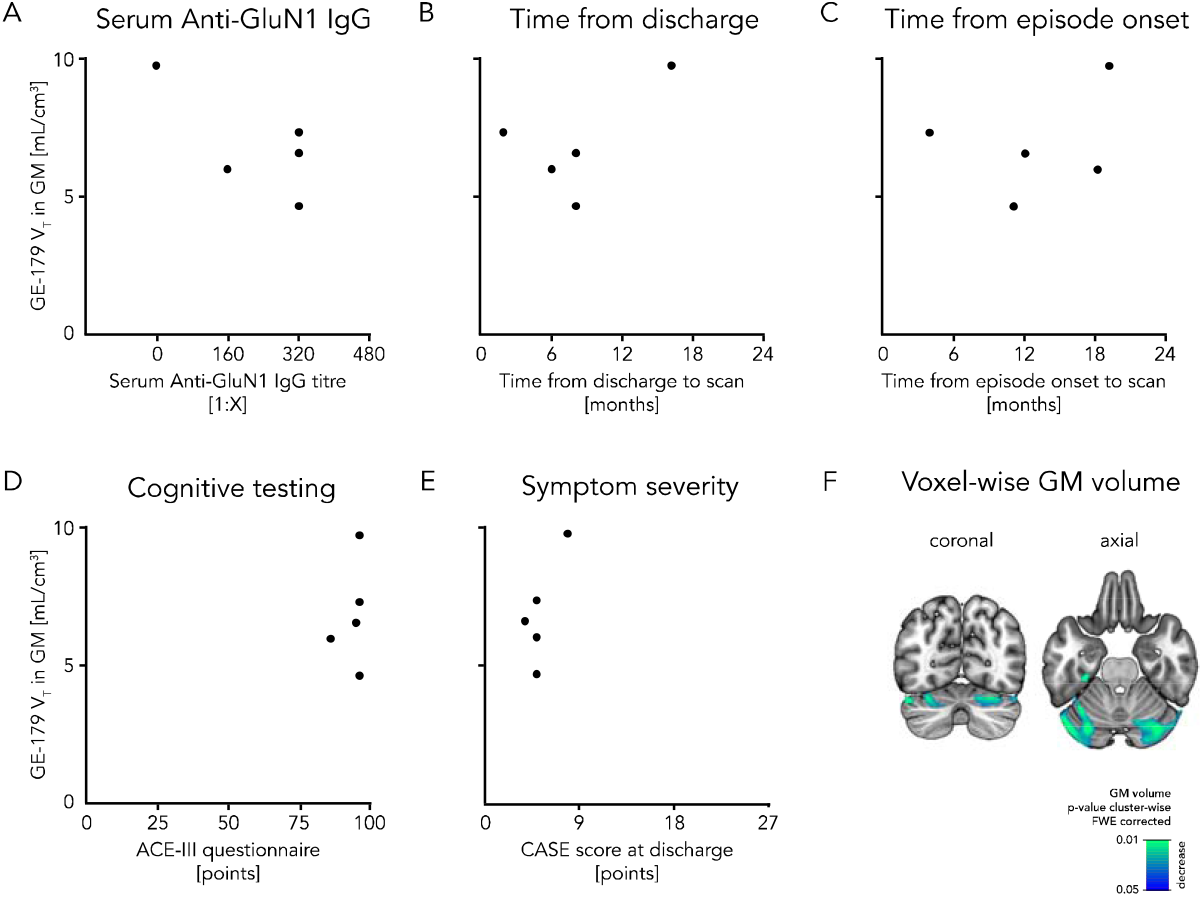
Correlation of [^18^F]GE-179 uptake with patient characteristics and MRI-based voxel-based morphometry. Scatter plots (**panels A to E**) show the association of [^18^F]GE-179 total volume of distribution (V_T_) in NMDAR-antibody encephalitis cases (n=5, both “seropositive” and “seroreverted” cases) with serum GluN1-specific immunoglobulin G (IgG, **panel A**), time from discharge to scan (**panel B**), time from episode onset to scan (**panel C**), cognitive testing with Addenbrooke’s Cognitive Examination III (ACE-III, **panel D**), and symptom severity at discharge measured with the CASE score (**panel E**). **Panel F** displays voxel-wise differences in grey matter volume between encephalitis cases and healthy volunteers (p<0.05 family wise error [FWE] corrected, blue colours indicate atrophy in encephalitis cases).

### Gray matter atrophy and neurotransmitter levels assessed using MRI

We assessed grey matter atrophy using MRI-based voxel-based morphometry (VBM). We found reduced grey matter volume in the cerebellar hemispheres (left, T = 4.6, 7388 mm^3^, p_FWE_ = 0.001; right, T = 4.3, 4512 mm^3^ voxels, p_FWE_ = 0.008; **Fig. 3F**). There was no overlap between areas of decreased grey matter volume and regions with significantly reduced [^18^F]GE-179 V_T_ (**Fig. 2B)**.

For all subjects imaged at UCL, we evaluated whether changes in radiotracer uptake correlated with hippocampal neurotransmitter levels, as assessed by the glutamate plus glutamine (Glx) to total creatine (Glx:tCr) ratio from magnetic resonance spectroscopy (MRS). The Glx:tCr ratio was no different between seropositive cases and healthy controls (2.6 [95% CI 0.5 – 4.6] vs. 1.5 [95% CI 0.5 – 2.4], F=1.0, p=0.35) but Glx:tCr ratio in the hippocampi negatively correlated with [^18^F]GE-179 uptake in grey matter (Pearson’s partial correlation coefficient -0.63, p=0.04).

## DISCUSSION

In this study, we report the first direct *in vivo* measure of activation of NMDA receptors in patients with definite NMDAR-antibody encephalitis. We demonstrated a regional reduction in the density of open, active NMDA receptors, most prominently in the anterior temporal and superior parietal cortices, in a small series of patients with persisting serum GluN1-autoantibodies. In contrast, a clinically completely recovered patient with seroreversion of GluN1-autoantibodies had a normal density of open, active NMDA receptors.

These human-derived results support the cross-linking and internalisation of NMDARs by GluN1-autoantibodies, as observed *in vitro*, in rodent studies and in *post mortem* specimens *(6, 8, 9)*. In our study, we observed a large reduction in active (open) NMDA receptors (mean 30% grey matter reduction in persistently seropositive patients compared to healthy volunteers) two to eight months after hospital discharge when there were mild residual symptoms in the studied patients. This points to the considerable compensatory capacity of the human brain.

NMDA receptors subserve long-term potentiation *(17)*, a process relevant for memory consolidation. Sustained activation of NMDA receptors is relevant for persistent neuronal activity that is the basis for storage of working memory *(18, 19)*. Impaired memory and executive function are common following NMDAR-antibody encephalitis *(5)*. Our results suggest that a persisting disruption of NMDAR function could be a potential cause of these deficits.

The abnormal density of active (open) NMDA receptors in patients with NMDAR-antibody encephalitis was not distributed uniformly throughout the cortex. The density of active NMDA receptors was mostly reduced in the anterior temporal lobes, which are important for episodic memory *(20)*, and parietal / posterior cingulate cortices, that are involved in psychosis *(21, 22)*. These regional alterations in NMDAR function may help explain the spectrum of symptoms observed in NMDAR-antibody encephalitis. The explanation for these regional differences, however, remains unclear.

We did not find significant correlations of [^18^F]GE-179 V_T_ in grey matter with patient characteristics. A small sample size and little variability between patients are potential reasons for a lack of significant correlations. Future studies may assess larger and more heterogenous cohorts and utilize more sensitive cognitive testing to uncover potential associations of [^18^F]GE-179 uptake with clinical variables.

We used MRS to assess neurotransmitter disturbances in the hippocampi of the included patients. There were no absolute differences of Glx:tCr ratios between patients and controls. However, higher Glx:tCr correlated with lower [^18^F]GE-179 uptake in grey matter. This may reflect increased neurotransmitter release in patients with more severely impaired NMDAR function due to receptor cross-linking and internalisation.

Our results of altered density of active NMDA receptors were not explained by brain atrophy because V_T_ values were partial-volume corrected. MRI-based VBM only showed atrophy in the cerebellar hemispheres and the statistical map did not overlap with abnormal NMDAR density observed on PET. The findings were also independent of brain perfusion, because V_T_ estimates are corrected for local and global blood perfusion effects.

Limitations of our study include a small sample size, and a different age and sex distribution of healthy volunteers and patients. The healthy control group was obtained by pooling [^18^F]GE-179 PET data from different studies. These cohorts rarely included females of reproductive age to reduce the potential risks of radiation. To minimise the influence of confounds on between-group comparisons, we corrected all analyses for age, sex, and site. Additionally, there was no association of scanning site (i.e. scanning equipment) with grey matter V_T_ (F=1.2, p=0.95). All female healthy volunteers younger than 40 years had V_T_ estimates in grey matter above 8 (mean 9.3, 95% CI 6.3-12.3), compared to 6.2 (95% CI 4.4 – 8.0) in seropositive cases, supporting the relevance of our results despite the between-cohort differences in sex and age.

We could only scan patients who tolerated a 70-minute PET scan and selected those not taking antipsychotics or antidepressants to prevent our results being confounded by movement artefacts or the effects of medication on NMDA receptors. We did not include cases with severe symptoms or anesthetised patients, because anaesthetics are likely to reduce the baseline activity of NMDA receptors *(15)*. Finally, we did not measure intrathecal GluN1-autoantibody titres, which are a better marker of disease activity than serum titres, as cerebrospinal fluid could not be obtained on the day of scanning due to ethical considerations.

If validated in larger and longitudinal studies, [^18^F]GE-179 PET could have the potential for tracking NMDAR function -the key predicted feature of disease activity - and guide accurate prognostication and escalation of immunotherapies or administration of NMDAR-active agents *(23)*, in addition to providing novel insights into the principle biological disease mechanism.

## MATERIALS AND METHODS

### Study population

We screened consecutive patients with NMDAR-antibody encephalitis that were hospitalised or under regular follow-up in two tertiary referral centres in the UK (John Radcliffe Hospital, Oxford University Hospitals NHS Foundation Trust, and St. George’s University Hospital NHS Foundation Trust London). We included patients with definite disease according to established diagnostic criteria *(13)* and positive GluN1-autoantibodies in CSF at diagnosis who were (i) between 18 and 65 years old; (ii) had minimal or no symptoms and thus could tolerate a 70-minute PET-MR scan; and (iii) did not take any medication that could interfere with NMDA receptors, in particular antipsychotics or antidepressants, permitting immunotherapy and anticonvulsants. The included patients were divided into a group scanned two to eight months after hospital discharge with mild symptoms and persistent “seropositivity” (cases #1-4) that had detectable GluN1-autoantibodies in serum on the day of scanning, and a “seroreverted” participant (case #5) that did not have detectable GluN1-autoantibodies in serum on the day of scanning, was scanned > 1 year after hospital discharge and was completely symptom free. All patients were scanned at University College London (UCL site), UK. Clinical data were retrospectively extracted from medical records according to previously described procedures *(1)*. We calculated the CASE score, a measure of autoimmune encephalitis severity, at discharge (range 0 to 27 points; higher scores indicate more severe disease) *(24)*. Cognitive testing with Addenbrooke’s Cognitive Examination III (ACE-III; range 0 to 100 points; higher scores indicate better cognitive function; scores > 89 typically considered normal) and assessment with Beck’s Depression Inventory (BDI; range 0 to 63 points; higher scores indicate more depressive symptoms; scores < 10 typically considered normal) were performed shortly before the scan.

We also included 29 healthy volunteers that had a [^18^F]GE-179 PET scan at one of three UK sites: 10 at the UCL site; 10 at Addenbrooke’s Hospital in Cambridge (Cambridge site); and 9 at Hammersmith Hospital in London (Hammersmith site). Healthy volunteers did not have any relevant neurological or psychiatric disorders and were not taking any regular medication.

The study was approved by the local research ethics committee and the local National Health Service Trust research office. All participants gave written informed consent.

### Data acquisition

All participants underwent dynamic emission PET scans after intravenous injection of a target dose of 185 MBq of [^18^F]GE-179. [^18^F]GE-179 binds specifically to the phencyclidine site inside the NMDA receptor channel *(25)*. Tracer binding requires the opening of the receptor channel complex. Successful blocking experiments were performed *in vivo* for GE-179 in rats following activation of NMDA receptors using electrical stimulation *(14)* and for the structural analogues CNS-5161 and GMOM in non-anaesthetised rodents *(26, 27)*, confirming the specificity of the tracer to the phencyclidine site within the open NMDA receptor ion channel. Because anaesthesia is expected to reduce the baseline opening probability of NMDA receptors, blocking experiments in anaesthetized rodents were not successful *(15)*.

The UCL site used a Siemens Biograph mMR combined PET-MR scanner for acquisition of 70-minute dynamic emission scans with a 25.8 cm axial field of view. To make the scanning procedure more easily applicable in clinical practice, as requested by the study funder (Medical Research Council), at the UCL site the scan duration was shortened from 90 to 70 minutes and an image-derived input function (IDIF) was used instead of arterial sampling.

Our group showed that shortening the scan duration to 70 minutes provides V_T_ estimates that correlate excellently (Spearman’s rank correlation coefficient 0.99, 95% CI 0.98 - 0.99, p<0.001) with those obtained from 90 minute scans and the results were robust for all studied subregions (7 distributed regions, Spearman’s rank correlation coefficient ranging from 0.98 to 1.00) *(28)*. List-mode data were initially processed using an in-house motion detection algorithm based on a principle component analysis of the dynamic PET signal *(29)*. Frame timing was individually adapted to exclude the typically short (duration between seconds to max. 1 min) signal alterations and reduce the effect of interframe sudden head movement.

This resulted in a variable number of frames (45 to 54). Data were reconstructed with 3-dimensional filtered backprojection to voxel sizes of 1.40 × 1.40 × 2.03 mm. Venous blood samples were obtained 7, 12, 22, 42, and 62 minutes after [^18^F]GE-179 injection to provide plasma-to-whole blood and parent fraction corrections to the IDIF *(30)*. The MR protocol included a high-resolution volumetric T1-weighted sequence (MPRAGE) with 2000 ms repetition time, 2.92 ms echo time, 256 × 256 matrix, and 1.1 × 1.1 × 1.1 mm voxels. To facilitate the IDIF we also performed an arterial time-of-flight magnetic resonance angiography (TOF-MRA) sequence that included the carotid arteries with 22ms repetition time, 4.17ms echo time, 256 × 256 matrix, and 0.625 × 0.625 × 2mm voxels. The magnetic resonance spectroscopy (MRS) acquisition (PRESS, repetition time, 2000ms; echo time 30ms) involved the manual placement and estimation of a single voxel covering the hippocampi (left and right separately) of each subject with approximate dimensions 30 mm (Anterior—Posterior), 16 mm (Left—Right), and 16 mm (thickness). One-hundred-twenty transients were collected per measurement.

The Cambridge site used a GE Discovery 690 combined PET-CT scanner for acquisition of 90-minute dynamic emission scans with a 15.4 cm axial field of view. Data were reconstructed with 3-dimensional filtered backprojection to 58 frames with voxel sizes of 2.00 × 2.00 × 3.27 mm. Arterial blood from the radial artery was continuously sampled for the first 6.5 minutes of the scan with discrete arterial samples taken at 0.5, 4.5 10, 15 and 20 minutes post-injection, and at 10-minute intervals thereafter. Each volunteer underwent 3T brain MRI on a Siemens Trio or Skyra system (Siemens Healthineers, Erlangen, Germany) including a high-resolution volumetric T1-weighted sequence (MPRAGE).

The Hammersmith site used a Siemens/CTI ECAT EXACT HR+ model 962 PET scanner for acquisition of 90-minute dynamic emission scans with a 15.5 cm axial field of view *(25)*.

Data were reconstructed with Fourier rebinning combined with 2-dimensional filtered back-projection to 34 frames with voxel sizes of 2.09 × 2.09 × 2.42 mm. Arterial samples were obtained continuously for the first 15 minutes, and discretely at baseline and at eight further times throughout the remainder of the scan. Each volunteer underwent 3T MRI on a GE Signa HDx system acquiring a T1-weighted coronal SPGR sequence with a voxel size of 0.938 mm × 1.100 mm × 0.938 mm.

### Data preprocessing

We performed data pre-processing following a previously described and validated protocol *(30)*. The Cambridge and Hammersmith sites used arterial sampling, with the input function corrected by plasma parent fraction and plasma-to-whole blood ratio determined from the arterial samples. The UCL site used an IDIF from the carotid arteries corrected by plasma parent fraction and plasma-to-whole blood ratio estimated from venous samples. The IDIF involved segmentation of the carotid arteries on TOF-MRA scans followed by extraction and partial volume correction of the radioactivity concentration in these arteries from the dynamic PET data. We previously validated this method by showing that the combination of an IDIF with venous samples provides reliable [^18^F]GE-179 V_T_ estimates compared to arterial sampling (Pearson’s correlation coefficient 0.95, p<0.001) *(30)*.

We fitted arterial (Cambridge and Hammersmith sites) and venous (UCL site) radioactivity concentrations, plasma parent fractions, and plasma-to-whole-blood ratios as described previously *(30)*. Site-specific spatial smoothing was applied to adjust the PET images from all sites to the same spatial resolution. Dynamic PET images were corrected for head motion using a post-hoc frame-to-frame realignment method and the realigned PET images were linearly co-registered with the T1-weighted MRI using Statistical Parametric Mapping 12 (SPM12, Wellcome Centre for Human Neuroimaging, London, https://www.fil.ion.ucl.ac.uk/spm/software/spm12/). We parcellated brain structures on T1-weighted MRIs using an algorithm based on Geodesic Information Flows (GIF) *(31)*, and adjusted the PET images for partial volume effects using the iterative Yang method *(32)* implemented in the PET-PVC toolbox *(33)*. Regional partial volume-corrected time-activity curves were extracted and modelled using a 2-brain-compartment 4-rate-constant kinetic model with a variable blood volume component as described previously *(25)*. Total V_T_ in mL/cm^3^ was estimated for overall grey and white matter and lobar regions of interest as a metric of radiotracer binding.

Additionally we computed V_T_ for each scan on a voxel level using Logan graphical analysis *(34)*. We used the Computational Anatomy Toolbox (CAT12, http://www.neuro.uni-jena.de/cat/) in SPM12 to segment and nonlinearly register all T1-weighted MRI images to a standard template. During this process we also extracted parametric grey matter volume images. We applied the same transformations to co-registered V_T_ images. We performed median filtering and gaussian smoothing with an 8 mm full-width-at-half-maximum kernel. For all subjects imaged at the UCL site, MRS data were phase and frequency corrected, before being summed into a single spectrum. A Hankel singular value decomposition filter was applied to remove the residual water signal and metabolites were quantified using a simulated basis set in Tarquin (http://tarquin.sourceforge.net). The average of the glutamate plus glutamine (Glx) to total creatine (Glx:tCr) ratio in the left and right hippocampus was extracted for each subject.

The live cell-based assays for serum GluN1-autoantibody titre measurements were performed as follows. HEK293T cells were cultured at 37°C in Dulbecco’s Modified Eagle Medium (DMEM) supplemented with foetal calf serum and antibacterial/antimycotic solution for 30 hours on glass cover slips, then transfected with plasmid DNA containing GluN1 subunit for 15 hours. Sera were tested following incubation for a further 24 hours in 1.4 μL/ml ketamine supplemented culture medium. Serum, previously thawed to room temperature, was diluted 1:20 with DMEM supplemented with 1% bovine serum albumin and 200mM 4-(2-hydroxyethyl)-1-piperazineethanesulfonic acid (HEPES). 250μL diluted samples were tested alongside positive and negative controls. Samples were incubated with transfected cells for 1 hour at room temperature. The diluted samples were then aspirated, each well was washed 3 times with DMEM-HEPES, each cover slip was then fixed with 4% formaldehyde for 5 minutes and then washed three times with DMEM-HEPES. The cells were then incubated with secondary antibody (Alexa Fluor 594-conjugated donkey anti-human IgG Fc-gamma; Jackson 709-585-098) in the dark for 45 minutes. This was aspirated, and each well was washed twice in DMEM-HEPES then twice in PBS. Each coverslip was individually mounted in 4′,6-diamidino-2-phenylindole (DAPI)-supplemented mounting medium on a glass slide. Following review of positive and negative control cover slips, test samples were read at 40x zoom on a Leica DM2000 fluorescence microscope. Positivity was determined by the presence of a characteristic coronal cell surface deposition of fluorescent reporter antibody on multiple cells. All positive samples were repeated on an independent assay and serially diluted to establish an end-point dilution, the dilution at which the positive signal was still unambiguously present. The threshold for seropositivity was set at > 1:20.

### Statistical analysis

We compared regional V_T_ estimates and hippocampal MRS data between patients with NMDAR-antibody encephalitis and healthy volunteers using the general linear model adjusting for age, sex, and site (i.e. scanning equipment). We analysed the four “seropositive” patients and one “seroreverted” patient separately. We report V_T_ and Glx:tCr mean values as estimated marginal means that were adjusted for age, sex, and, for V_T_, site. We calculated the correlation between hippocampal MRS data and grey matter [^18^F]GE-179 V_T_ using Pearson’s partial correlation adjusting for sex and age.

To analyse the subregional distribution of the findings, we assessed the spatial distribution of differences between groups for voxel-wise V_T_ estimates. To address the contribution of grey matter atrophy to our findings, we performed voxel-based morphometry (VBM) using segmented, normalised and modulated parametric grey matter volume images. Both voxel-wise datasets were assessed using general linear models corrected for age, sex, and site. We report voxel-wise p-values at a threshold of <0.05 on a cluster-level family-wise error corrected for multiple comparisons (p_FWE_).

## Data Availability

All data produced in the present study are available upon reasonable request to the authors.

## Acknowledgements

The authors thank the staff at GE Healthcare, in particular William Trigg, Sajinder Kaur Luthra and Jo Stevens, for their help and support during this study.

This work was undertaken in part at UCL/UCLH which receives support from the NIHR University College London Hospitals Biomedical Research Centre. AH was supported by Medical Research Council Clinician Scientist Fellowship (G108/585) and MRC Clinical Sciences Centre core funding (MC_U120085812).

Authors affiliated with 9 acknowledge support by the UK Department of Health via the National Institute for Health Research (NIHR) comprehensive Biomedical Research Centre award to Guy’s & St Thomas’ NHS Foundation Trust in partnership with King’s College London and King’s College Hospital NHS Foundation Trust, and by the Wellcome EPSRC Centre for Medical Engineering at King’s College London (WT 203148/Z/16/Z).

During the period of the study AAD was the recipient of a Wellcome Trust clinical research training fellowship (205126/Z/16/Z), the 2017 British Medical Association (BMA) Foundation Margaret Temple grant, and supported by the National Institute for Health Research (NIHR) Oxford and NIHR Oxford Health Biomedical Research Centres. CM was supported by the Medical Research Council (MR/N013042/1) and subsequently by the Wellcome Trust/Engineering and Physical Sciences Research Council (EPSRC) Centre for Medical Engineering (WT 203148/Z/16/Z) and the Engineering and Physical Sciences Research Council Centre for Doctoral Training in Medical Imaging (EP/L015226/1). JPC & FIA report a UK Medical Research Council (MRC) grant (MRC Industry Collaboration Agreement (MR/K02308X/1)), and MK, JPC & FIA report a UK MRC grant (Developmental Pathway Funding Scheme (MR/L013215/1)). JPC reports a British Journal of Anaesthesia/Royal College of Anaesthetists grant from the National Institute of Academic Anaesthesia. JPC is supported by the Cambridge NIHR Biomedical Research Centre. The views expressed are those of the authors and not necessarily those of the NIHR or the Department of Health and Social Care.

## Author contributions

<u>Conception or design of the work:</u> MG, AAD, SRI, MJK

<u>Data collection:</u> AH, MG, CJM, SRI, AAD, JPC, KS, EÅ, UV, FIA, TDF, YTH

<u>Data analysis and interpretation:</u> MG, SRI, AAD, KS, EÅ, FT, CJS, AB, KE, BAT, KT, MCW

<u>Study coordination:</u> MJK, SRI, CJM, AH, JPC

<u>Drafting the article:</u> MG, AAD, UV, SRI, MCW, MJK

<u>Critical revision of the article:</u> all authors

