## Supplementary material for "*In vivo* NMDA receptor function in people with NMDA receptor antibody encephalitis": Online Supplement

### NEST Investigators consortium

| Author | Affiliations |
|---|---|
| *Franklin I Aigbirhio* | • Wolfson Brain Imaging Centre, Department of Clinical Neurosciences, University of Cambridge, Addenbrooke's Hospital, Cambridge, UK |
| *Adam Al-Diwani* | • Oxford Autoimmune Neurology Group, Nuffield Department of Clinical Neurosciences, University of Oxford, Oxford, United Kingdom<br>• Department of Psychiatry, University of Oxford, Oxford, United Kingdom |
| *Gareth Ambler* | • Department of Statistical Science, University College London, London, UK |
| *Erik Årstad* | • Centre for Radiopharmaceutical Chemistry, University College London, London, UK |
| *Anna Barnes* | • Institute of Nuclear Medicine, University College London Hospitals, London, UK |
| *Ilaria Boscolo Galazzo* | • Department of Computer Science, University of Verona, Verona, Italy |
| *Roberto Canales* | • Wolfson Brain Imaging Centre, Department of Clinical Neurosciences, University of Cambridge, Addenbrooke's Hospital, Cambridge, UK |
| *Sarah Chetcuti* | • Division of Anaesthesia, Department of Medicine, University of Cambridge, Addenbrooke's Hospital, Cambridge, UK |
| *Jonathan P Coles* | • Division of Anaesthesia, Department of Medicine, University of Cambridge, Addenbrooke's Hospital, Cambridge, UK |
| *Bianca De Blasi* | • Department of Medical Physics and Bioengineering, University College London, London, UK |
| *John C Dickson* | • Institute of Nuclear Medicine, University College London Hospitals, London, UK |
| *John S Duncan* | • Department of Clinical and Experimental Epilepsy, UCL Queen Square Institute of Neurology, London, UK<br>• MRI Unit, Chalfont Centre for Epilepsy, UK |
| *Simon Eaglestone* | • Translational Research Office, University College London, London, UK |
| *Evan Edmond* | • Physiological Neuroimaging Group, Nuffield Department of Clinical Neurosciences, University of Oxford, Oxford, United Kingdom |
| *Kjell Erlandsson* | • Institute of Nuclear Medicine, University College London Hospitals, London, UK |
| *Martin Fisher* | • Wolfson Brain Imaging Centre, Department of Clinical Neurosciences, University of Cambridge, Addenbrooke's Hospital, Cambridge, UK |

| | |
|---|---|
| *Tim D Fryer* | • Wolfson Brain Imaging Centre, Department of Clinical Neurosciences, University of Cambridge, Addenbrooke's Hospital, Cambridge, UK |
| *Marian Galovic* | • Department of Neurology, Clinical Neuroscience Center, University Hospital Zurich, Zurich, Switzerland<br>• Department of Clinical and Experimental Epilepsy, UCL Queen Square Institute of Neurology, London, UK<br>• MRI Unit, Chalfont Centre for Epilepsy, UK |
| *Thibault Gendron* | • Centre for Radiopharmaceutical Chemistry, University College London, London, UK |
| *Ashley Groves* | • Institute of Nuclear Medicine, University College London Hospitals, London, UK |
| *Alexander Hammers* | • King's College London and Guy's and St Thomas' PET Centre, Division of Imaging Sciences & Biomedical Engineering, King's College London, London, UK |
| *Abigail Hancock-Scott* | • Wolfson Brain Imaging Centre, Department of Clinical Neurosciences, University of Cambridge, Addenbrooke's Hospital, Cambridge, UK |
| *Young T Hong* | • Wolfson Brain Imaging Centre, Department of Clinical Neurosciences, University of Cambridge, Addenbrooke's Hospital, Cambridge, UK |
| *Brian Hutton* | • Institute of Nuclear Medicine, University College London Hospitals, London, UK |
| *Sarosh R Irani* | • Oxford Autoimmune Neurology Group, Nuffield Department of Clinical Neurosciences, University of Oxford, Oxford, United Kingdom<br>• Department of Neurology, John Radcliffe Hospital, Oxford University Hospitals NHS Foundation Trust, Oxford, UK |
| *Ilijas Jelcic* | • Department of Neurology, Clinical Neuroscience Center, University Hospital Zurich, Zurich, Switzerland |
| *Matthias J Koepp* | • Department of Clinical and Experimental Epilepsy, UCL Queen Square Institute of Neurology, London, UK<br>• MRI Unit, Chalfont Centre for Epilepsy, UK |
| *Roido Manavaki* | • Department of Radiology, University of Cambridge, Addenbrooke's Hospital, Cambridge, UK |
| *Colm J McGinnity* | • King's College London and Guy's and St Thomas' PET Centre, Division of Imaging Sciences & Biomedical Engineering, King's College London, London, UK |
| *Lindsay McMurray* | • Wolfson Brain Imaging Centre, Department of Clinical Neurosciences, University of Cambridge, Addenbrooke's Hospital, Cambridge, UK |
| *Sarah McQuaid* | • Institute of Nuclear Medicine, University College London Hospitals, London, UK |

| | |
|---|---|
| *Alaleh Rashidnasab* | • Institute of Nuclear Medicine, University College London Hospitals, London, UK |
| *Joseph J Russell* | • Wolfson Brain Imaging Centre, Department of Clinical Neurosciences, University of Cambridge, Addenbrooke's Hospital, Cambridge, UK |
| *Kerstin Sander* | • Centre for Radiopharmaceutical Chemistry, University College London, London, UK |
| *Hasan Sari* | • Institute of Nuclear Medicine, University College London Hospitals, London, UK<br>• Athinoula A. Martinos Center for Biomedical Imaging, Department of Radiology, Massachusetts General Hospital and Harvard Medical School, Charlestown, MA, USA |
| *Selena Sephton* | • Wolfson Brain Imaging Centre, Department of Clinical Neurosciences, University of Cambridge, Addenbrooke's Hospital, Cambridge, UK |
| *Daichi Sone* | • Department of Clinical and Experimental Epilepsy, UCL Queen Square Institute of Neurology, London, UK<br>• MRI Unit, Chalfont Centre for Epilepsy, UK |
| *Charlotte J Stagg* | • Physiological Neuroimaging Group, Nuffield Department of Clinical Neurosciences, University of Oxford, Oxford, United Kingdom |
| *Kris Thielemans* | • Institute of Nuclear Medicine, University College London Hospitals, London, UK |
| *Benjamin A Thomas* | • Institute of Nuclear Medicine, University College London Hospitals, London, UK |
| *Francisco Torrealdea* | • Department of Medical Physics and Biomedical Engineering, University College London Hospitals, London, UK |
| *Umesh Vivekananda* | • Department of Clinical and Experimental Epilepsy, UCL Queen Square Institute of Neurology, University College London, London, UK |
| *Matthew C Walker* | • Department of Clinical and Experimental Epilepsy, UCL Queen Square Institute of Neurology, University College London, London, UK |
